# Adversarial Robustness of Capsule Networks for Medical Image Classification

**DOI:** 10.64898/2026.03.09.26347900

**Authors:** Anand Srinivasan, Durga V. Sritharan, Saahil Chadha, Daniel Fu, Jahid Omar Hossain, Gregory A. Breuer, Sanjay Aneja

## Abstract

**Purpose:** Deep learning models are increasingly being used in medical diagnostics, but their vulnerability to adversarial perturbations raises concerns about their reliability in clinical applications. Capsule networks (CapsNets) are a promising architecture for medical imaging tasks, given their ability to model spatial relationships and train with smaller amounts of data. Although previous studies have focused on adversarial training approaches to improve robustness, exploring alternative architectures is an underexplored direction for combating poor adversarial stability. Prior work has suggested that CapsNets may exhibit improved robustness to adversarial perturbations compared to convolutional neural networks (CNNs), but performance on adversarial images has not been studied systematically in clinical environments. We evaluated the robustness of CapsNets compared to CNNs and vision transformers (ViTs) across multiple medical image classification tasks.

**Methods:** We trained two CNNs (ResNet-18 and ResNet-50), one ViT (MedViT), and two CapsNets (DR-CapsNet and BP-CapsNet) on four distinct medical imaging datasets (PneumoniaMNIST, BreastMNIST, NoduleMNIST3D, and BloodMNIST) and one natural image dataset (MNIST). Models were evaluated on adversarial examples generated by projected gradient descent, fast gradient sign method, AutoAttack, Carlini & Wagner, and black-box transfer attacks. Models also underwent PGD adversarial fine-tuning to assess whether architectural differences persisted after adversarial training. Interpretability experiments, including latent space and Gradient-weighted Class Activation Mapping (Grad-CAM) analyses, were conducted to better understand model stability on adversarial inputs.

**Results:** Under conventional training, CapsNets demonstrated superior robustness under adversarial perturbations compared to CNNs and ViTs across all medical imaging datasets and the natural image dataset. Latent space and Grad-CAM visualizations revealed that CapsNets maintained more consistent feature representations and attention maps after adversarial perturbations compared to CNNs and ViTs, suggesting that advantages in CapsNet robustness are supported, at least in part, by more stable feature encodings. PGD adversarial fine-tuning improved robustness across models, with MedViT exhibiting the largest gains and thereby narrowing the performance gap.

**Conclusion:** CapsNets exhibit advantages in adversarial robustness over CNN- and ViT-based models on medical imaging tasks. These findings suggest that capsule-based architectures may be a promising direction for robust medical image classification. Future work should validate CapsNet robustness across more complex medical imaging tasks, independent external datasets, and broader sources of real-world imaging variability.

## Introduction

Deep learning (DL) has transformed diagnostic image analysis across multiple disease domains.^1–3^ DL methods have improved diagnostic efficiency and accuracy by reducing inter-observer variability and accelerating overall image interpretation.^1,4–9^ Nevertheless, widespread adoption of DL into clinical medicine remains in its nascency.^10,11^ Clinical implementation of well-trained DL models into diagnostic imaging practice is expected to increase, but many concerns regarding model reliability remain.^12–15^ Ensuring that DL systems maintain stable and trustworthy performance in the presence of real-world variability is critical for their clinical deployment.

Convolutional neural networks (CNNs) and vision transformers (ViTs) are the most widely used and mature DL architectures that have been applied to medical imaging data.^16,17^ Although CNN and ViT models have shown high fidelity across a range of diagnostic tasks, some studies have highlighted limitations in generalizability and stability.^18–23^ One notable concern with respect to stability is poor model performance on adversarial images. Adversarial images are images that have undergone targeted perturbations designed to mislead DL models, often resulting in incorrect predictions.^22–24^ The perturbations are often imperceptible to the human eye, and prior work has shown that altering as little as a single pixel can cause misclassification.^25^ Concerningly, medical DL algorithms have been found to exhibit an even higher sensitivity to adversarial images than non-medical algorithms, potentially due to complex biological textures that heighten vulnerability.^26,27^ Poor performance on adversarial images can be a strong indicator of a lack of model stability.^28–31^ Furthermore, adversarial examples call into question the real world reliability of medical DL models. Although adversarial perturbations are algorithmically constructed, they have been shown to expose model vulnerabilities that overlap with real-world sources of variability, including image noise, acquisition artifacts, and scanner heterogeneity. As such, adversarial robustness can serve as a useful stress test for model stability under clinically relevant distribution shifts. Adversarial robustness is therefore an important metric for evaluating medical DL models and ensuring their safe clinical implementation.

Prior efforts to improve robustness have largely focused on adversarial training strategies, which augment training data with perturbed examples. While these approaches can improve performance against specific attack types, they often provide limited generalization, introduce trade-offs with baseline accuracy, and may not fully address broader issues of model instability.^24,29,30,31^ One underexplored direction in creating stable DL imaging models is employing alternative architectures that confer inherent robustness to perturbations. Capsule networks (CapsNets) represent on such architecture and were designed to address the limitations of CNNs in modeling hierarchical relationships.^35^ In a CapsNet, neurons are divided amongst capsules, which compute vector outputs that convey both pose and probability information. Between capsule layers, a routing algorithm matches vector outputs from the preceding layer to the most applicable capsules in the subsequent layer. These properties may promote more stable internal representations and improved generalization, particularly in settings with limited data, making CapsNets an attractive candidate for medical imaging applications.^35–40^ To that end, CapsNets have shown strong performance across a range of medical tasks including COVID-19 detection, electrocardiogram classification, and brain image segmentation.^38–45^

Although CapsNets have demonstrated promising performance across a variety of medical imaging tasks, their robustness to adversarial perturbations has not been systematically evaluated in clinically relevant imaging settings.^44,45^ Evaluations of the adversarial performance of CapsNets on non-medical imaging tasks have yielded mixed results.^44–46^ Additionally, prior work has not compared the adversarial performance of CapsNets to more recently developed ViTs. Furthermore, the mechanisms underlying potential differences in robustness across architectures are not well understood, particularly with respect to the stability of learned feature representations.

In this study, we tested classification models against two common adversarial perturbation methods, comparing CapsNets to CNNs and ViTs across multiple medical imaging tasks spanning diverse modalities and classification settings. We assess model performance under varying strengths of adversarial perturbation and complement these analyses with interpretability experiments, including latent space and gradient-based visualization methods, to characterize the stability of internal representations. We further investigate a modified CapsNet architecture incorporating Bayes–Pearson routing to examine the impact of the capsule routing algorithm on robustness. Through this work, we aim to clarify the role of model architecture in promoting robustness and to identify design principles that may support the development of more reliable and clinically deployable AI systems.

## Methods

### Imaging Datasets

Four medical image datasets (PneumoniaMNIST, BreastMNIST, NoduleMNIST3D, and BloodMNIST) were selected to represent common medical image applications (Table 1).^47^ These datasets were chosen to capture a diverse range of medical imaging modalities (X-ray, ultrasound, computed tomography, and histology), imaging dimensions (2D and 3D), and classification tasks (binary and multi-class). As a non-medical image control group, MNIST was also included to contextualize model behavior outside of the medical domain. Spatial resolution was consistent across all datasets, with 2D images at a resolution of 28×28 pixels and 3D images at a resolution of 28×28×28 voxels.

**Table 1.** Characteristics of natural imaging (MNIST) and medical imaging (MedMNIST) datasets.

|  |  |  |  |
|---|---|---|---|
| PneumoniaMNIST | Chest X-Ray | 2 | 5,856 |
| BreastMNIST | Breast Ultrasound | 2 | 780 |
| NoduleMNIST3D | Chest Computed Tomography | 2 | 1,633 |
| BloodMNIST | Blood Cell Microscope | 8 | 17,092 |
| MNIST | Handwritten Digit Images | 10 | 70,000 |

### PneumoniaMNIST

PneumoniaMNIST (n=5,856; train/val/test = 4,708/524/624) is a collection of pediatric chest X-ray images. Models were trained to perform a binary classification task for pneumonia detection. The negative class corresponded to normal findings, and the positive class corresponded to the presence of bacterial or viral pneumonia.^48^

### BreastMNIST

BreastMNIST (n=780; train/val/test = 546/78/156) is a collection of breast ultrasound images. Models were trained to perform a binary classification task, where the negative class corresponded to normal or benign findings, and the positive class corresponded to malignant tumors.^49^

### NoduleMNIST3D

NoduleMNIST3D (n=1,633; train/val/test = 1,158/165/310) is a collection of 3D chest computed tomography volumes. Models were trained to perform a binary classification task, where the negative class corresponded to benign findings, and the positive class corresponded to malignant nodules, based on radiologist-assigned malignancy scores.^50^

### BloodMNIST

BloodMNIST (n=17,092; train/val/test = 11,959/1,712/3,421) is a collection of blood cell microscopy images. Models were trained to perform a multi-class classification task for blood cell type categorization. Labels corresponded to the following blood cell types: neutrophils, eosinophils, basophils, lymphocytes, monocytes, platelets, erythroblasts, and immature granulocytes.^51^

### Modified National Institute of Standards and Technology (MNIST)

MNIST (n=70,000; train/val/test = 52,800/7,200/10,000) is a collection of handwritten digits, widely used for benchmarking machine learning models. Models were trained to perform a multi-class classification task, with labels corresponding to the integers 0-9. The MNIST dataset was included as a control for all experiments evaluating adversarial robustness to ensure that models behaved as expected on a simple, well-characterized natural image benchmark.

### Deep Learning Computer Vision Models

Two CapsNet models, dynamic routing CapsNet (DR-CapsNet) and Bayes-Pearson CapsNet (BP-CapsNet), were trained on all datasets (Figure 1A). Both models have previously shown promise for medical image classification.^52,53^ DR-CapsNet follows the original CapsNet architecture proposed by Sabour et al.,^35^ consisting of an initial convolutional layer, a primary capsule layer, and a final capsule layer. Dynamic routing is used between the capsule layers to iteratively match capsule outputs in the prior layer to appropriate capsules in the following one. BP-CapsNet follows the same architecture but substitutes dynamic routing with a Bayes-Pearson routing algorithm.^52^ Bayes-Pearson routing replaces the dot product in the dynamic routing algorithm with a Pearson correlation coefficient to achieve a more precise similarity measure. A self-exclusion mechanism is also added to restrain the influence of noisy or weakly correlated capsule outputs.^52^ Both CapsNets use fewer parameters than conventional CNNs and have been shown to generalize well to new images, even with limited training data (Supplementary Table 1).^35–40^

**Fig. 1.**
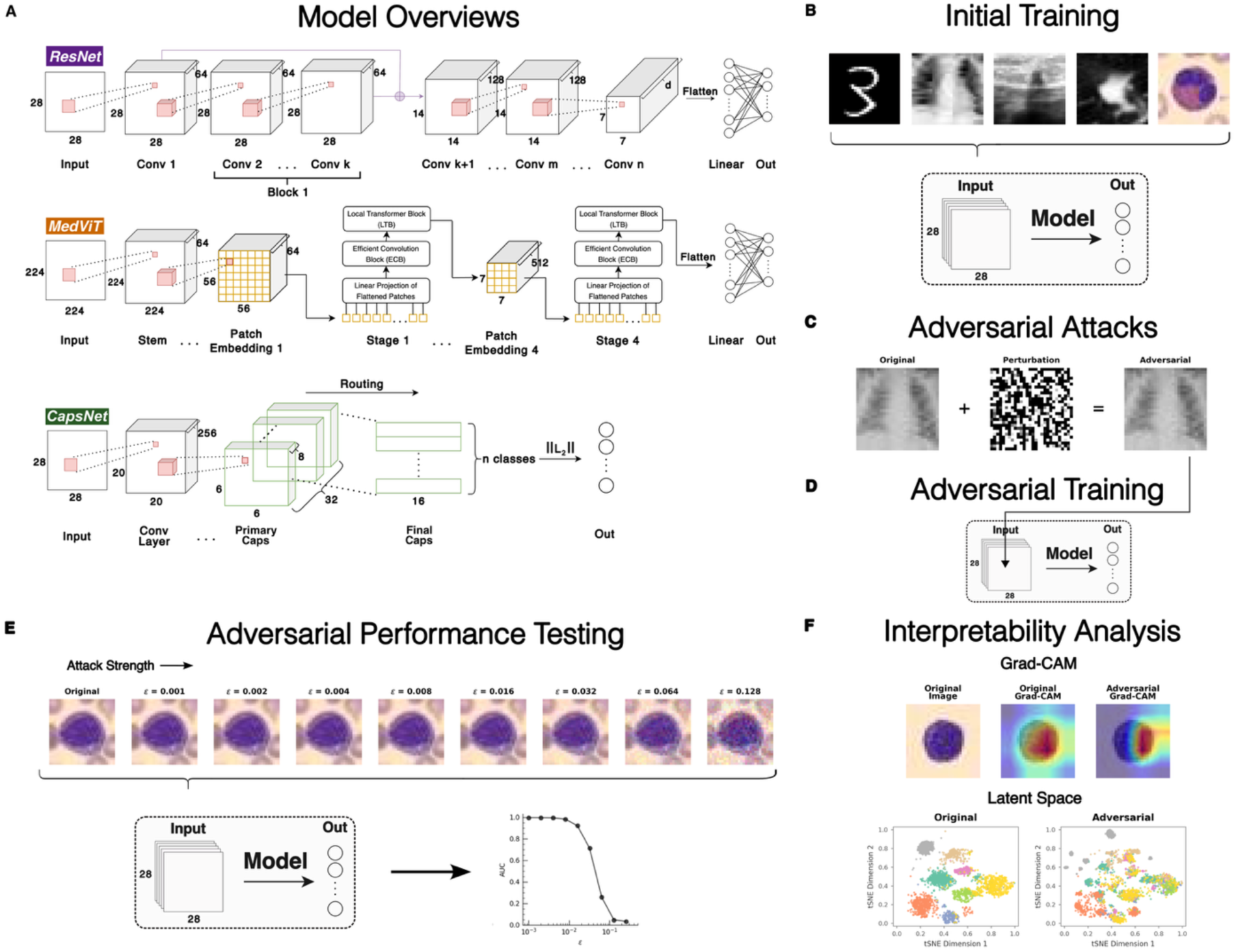
Overview of overall project workflow. **A.** Generalized residual network (ResNet), medical vision transformer (MedViT), and capsule network (CapsNet) model architecture diagrams. Five classification models—two ResNets (ResNet-18 and ResNet-50), one MedViT (MedViT-S), and two CapsNets (DR-CapsNet and BP-CapsNet)—were selected for analysis. **B.** All models were initially trained on one natural imaging dataset (MNIST) and four medical imaging datasets (PneumoniaMNIST, BreastMNIST, NoduleMNIST3D, and BloodMNIST). **C.** Adversarial image examples were generated through multiple methods, including projected gradient descent (PGD), fast gradient sign method (FGSM), AutoAttack, and Carlini & Wagner. **D.** Models underwent adversarial fine-tuning using PGD adversarial examples as input. **E.** All traditionally and adversarially trained models were tested against adversarial images with varying strength across all five datasets. **F.** Model features were analyzed using Gradient-weighted Class Activation Mapping (Grad-CAM) and latent space visualization to better understand how different models responded to adversarial perturbations.

For comparison, two popular CNN models (ResNet-18 and ResNet-50) and a medical ViT (MedViT) model were trained. ResNet-18 and ResNet-50 are residual networks consisting of 18 and 50 layers organized into basic and bottleneck blocks, respectively.^54^ Both networks employ residual connections between blocks to facilitate gradient flow. MedViT is a hybrid ViT that integrates convolutional and transformer-based components to support generalized medical image classification tasks.^55^ Specifically, MedViT-S, the best-performing MedViT model, was chosen for analysis. For NoduleMNIST3D, the 28 slices of each 28 × 28 × 28 volume were treated as input channels to the 2D architectures, and each model’s input layer was modified to accept 28 channels.

Model training followed a similar approach to Lei et al.^52^ For training on medical datasets, all models were trained for 100 epochs using the Adam optimizer with an initial learning rate of 1e-4 and a batch size of 16. An exponential learning rate scheduler was applied with a decay factor of 0.96 per epoch. All CapsNet models were trained with margin and reconstruction loss for regularization, with reconstruction-loss weights of 5x10^-4^ and 0.5, respectively.^35^ Cross entropy loss was used for all other models. For MNIST, models were trained for 20 epochs, which was sufficient for convergence. All other optimization and loss settings remained unchanged.

### Adversarial Perturbation Methods

To study the adversarial vulnerability of all models, four adversarial perturbation methods were implemented: projected gradient descent (PGD), fast gradient sign method (FGSM), AutoAttack, and the Carlini & Wagner L_2_ attack (CW) (Figure 1C). PGD and FGSM were selected as established white-box adversarial attacks that have been widely employed and are among the most frequently evaluated attack methods in medical imaging research.^56,57^ Both PGD and FGSM are gradient-based methods that seek to solve an optimization problem in which the objective is to maximize classification error while minimizing the magnitude of perturbation applied to the original input. PGD is considered a stronger adversarial perturbation method than FGSM because it performs multiple iterative steps of gradient ascent. As a result, PGD is often regarded as a universal first-order adversary, capable of approximating the worst-case perturbation within a bounded region.^32^ FGSM, on the other hand, is computationally efficient but thought to provide a weaker perturbation. AutoAttack and CW were additionally included as complementary adversarial attacks to provide a more comprehensive evaluation of model robustness and assess whether the observed resistance to FGSM and PGD persisted across alternative attack strategies. All attacks were untargeted and optimized classification scores without including the capsule reconstruction loss. Black-box transfer attacks were additionally evaluated using adversarial examples generated against different source architectures.

### Projected Gradient Descent

The PGD algorithm performs multiple iterative steps of gradient ascent, with projections back onto the allowed perturbation set after each iteration (Figure 2). A predefined perturbation bound ε determines the maximum allowable pixel-level change to the input. PGD is formally defined as follows:

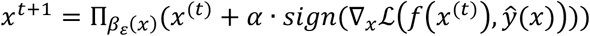

**Fig. 2.**
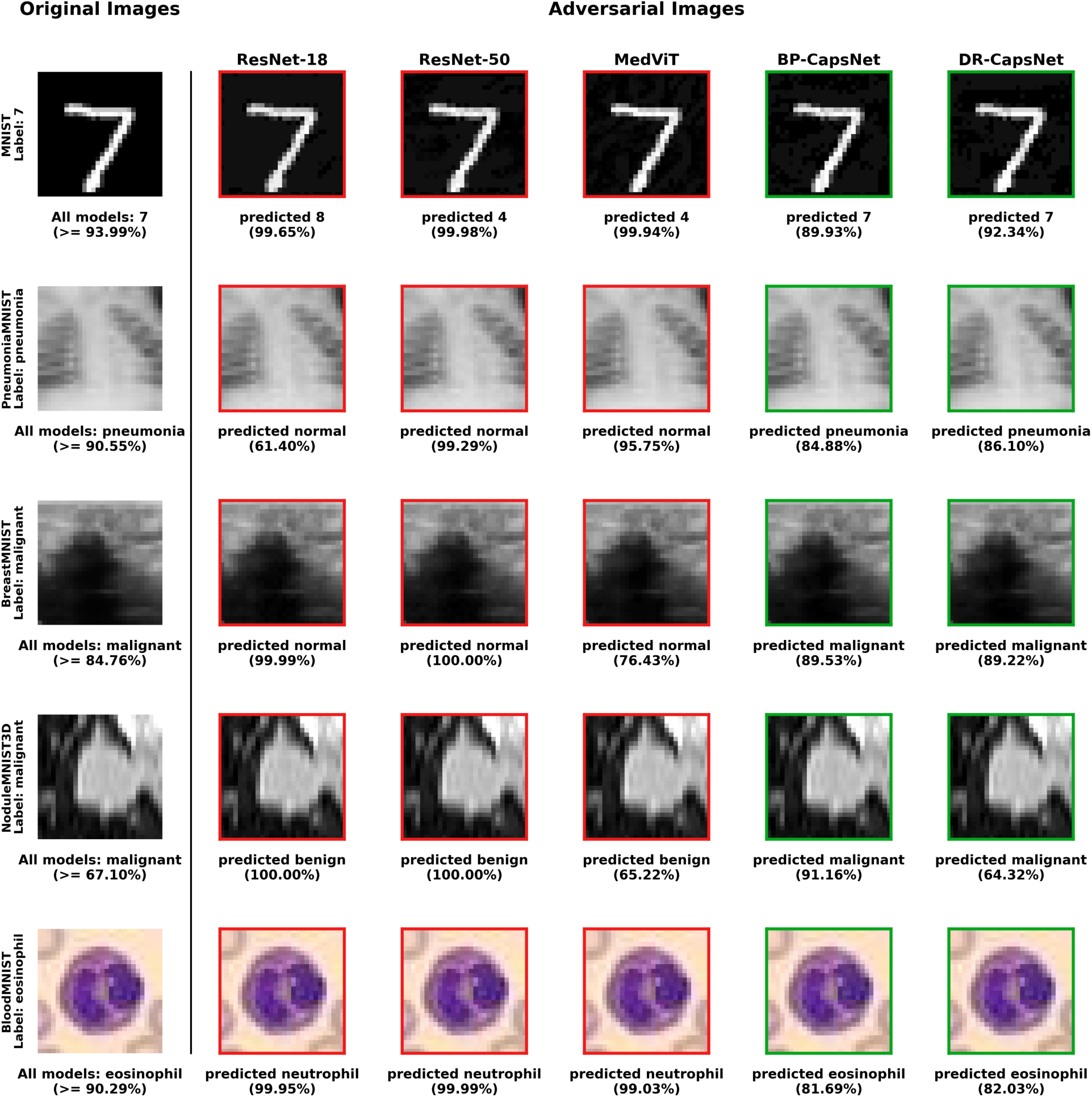
Examples of projected gradient descent (PGD) adversarial perturbations applied to natural and medical images. Adversarial image generation methods add targeted noise to input images. The resulting adversarial image examples can lead machine learning models to make incorrect classifications. Each adversarial image was generated using a PGD adversarial attack with an attack strength of *ɛ* = 0.128 for natural domain images and an attack strength of *ɛ* = 0.016 for medical domain images. All models correctly classified the original images, but the residual networks (ResNet-18 and ResNet-50) and the medical vision transformer (MedViT) failed to classify the perturbed examples. Model confidence for each prediction is shown below each image.

where *x*^(0)^ is the input, *x*^(*t*)^ is the adversarial example at iteration *t*, *ɛ* is the perturbation bound, *α* is the step size, *f*(*x*^(*t*)^0 is the model prediction, *ŷ*(*x*) is the model’s predicted class for the clean input x, ℒ is the loss function, ∇_*x*_ℒ is the gradient of the loss with respect to the input, and Π_*β_ɛ_* (*x*)_ is the projection onto the epsilon-ball.^32^

PGD was implemented as an untargeted *L*_∞_-constrained attack using the Adversarial Robustness Toolbox. For each perturbation bound, PGD was run for 30 iterations with step size *α*=*ɛ*/4 and three independent random initializations. Each initialization was generated by sampling a perturbation uniformly from the *L*_∞_ball of radius *ɛ* around the clean image. The attack maximized cross-entropy loss computed from the models’ raw classification scores; capsule reconstruction loss was excluded from the attack objective. As a sensitivity analysis, evaluation at *ɛ*=0.016 was repeated using 100 iterations and five random initializations.

### Fast Gradient Sign Method

FGSM is a simple, single-step perturbation method that perturbs an input image in the direction of the sign of the gradient. Similar to PGD, a perturbation bound ε defines the maximum allowable pixel-level change to the input. FGSM is formally defined as follows:

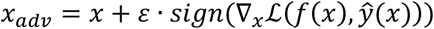

where *x* is the input, *x*_*adv*_ is the adversarial example, *ɛ* is the perturbation bound, *f*(*x*) is the model prediction, *ŷ*(*x*) is the model’s predicted class for the clean input x, ℒ is the loss function, and ∇_*x*_ℒ is the gradient of the loss with respect to the input.^22^

### AutoAttack

AutoAttack was implemented using the Adversarial Robustness Toolbox as an untargeted *L*_∞_ ensemble attack. For multiclass datasets, the ensemble consisted of Auto-PGD with cross-entropy loss, Auto-PGD with difference-of-logits-ratio loss, DeepFool, and the gradient-free Square Attack. Both Auto-PGD components used 100 iterations, five random initializations, and an initial step size of *ɛ* /4. DeepFool used 100 iterations with an overshoot parameter of 0.001, while Square Attack used 5,000 iterations, five restarts, and an initial fraction of perturbed features p_init_=0.8. Because difference-of-logits-ratio loss requires at least three classes, the binary-dataset ensemble excluded that component and consisted of Auto-PGD with cross-entropy loss, DeepFool, and Square Attack.

### Carlini & Wagner

The untargeted Carlini–Wagner L₂ attack was used to identify low-magnitude perturbations that changed the predicted class. Unlike PGD, FGSM, and AutoAttack, CW was not evaluated at fixed perturbation bounds. Instead, it jointly optimized perturbation magnitude and classification error using 500 optimization iterations, 10 binary-search steps, a learning rate of 0.01, an initial trade-off constant of c=0.01, and confidence parameter *κ*=0. CW was applied only to test examples that were initially classified correctly. Attack success rate (ASR) was calculated using initially correct examples as the denominator, and distortion statistics included successful attacks only. Because of the high computational cost of CW on BloodMNIST, models were evaluated on a common, reproducible, class-balanced cohort of 300 test images. All 300 images were correctly classified by all five architectures before attack.

For capsule models in which the initial c=0.01 configuration produced a low ASR, CW was repeated with an initial trade-off constant of c=10. These higher-c runs were performed as convergence analyses to determine whether apparent attack failures resulted from insufficient weighting of the classification objective.

Perturbation magnitudes were calculated in the model-input [0,1] pixel space. Because MedViT used 224×224 inputs whereas the other architectures used their native 28×28 inputs, raw L₂ norms were additionally converted to per-element root-mean-square (RMS) distortion:

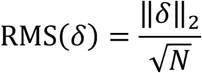

where *δ* is the adversarial perturbation and N is the number of input elements, including all channels or slices. RMS distortion was used for cross-architecture comparisons to account for differences in input dimensionality.

### Adversarial Robustness Testing

We evaluated the adversarial robustness of all five models across each dataset using complementary white-box, ensemble, optimization-based, and black-box transfer attacks. For the white-box PGD and FGSM evaluations, a model-specific adversarial test set was generated at each perturbation bound. Attacks were evaluated at *ɛ* ∈ {0.001, 0.002, 0.004, 0.008, 0.016, 0.032, 0.064, 0.128}. For MNIST, a fixed random subset of 1,000 images was sampled without replacement from the standard 10,000-image test set. The same clean-image subset was used across models and perturbation strengths, although adversarial examples were generated separately for each model. For all medical datasets, adversarial evaluation used the complete official test split. AutoAttack was used as a complementary ensemble evaluation on the four medical datasets, combining gradient-based and gradient-free attacks. Because of its substantially greater computational cost, AutoAttack was evaluated at a fixed, moderate perturbation bound *ɛ* =0.032, rather than over the complete epsilon sweep. Performance on each adversarial test set was evaluated using the metrics of area under the receiver operating characteristic curve (AUC) and accuracy. We would expect more robust models to maintain performance close to their baseline scores on the adversarial test sets, while less robust models would experience more rapid performance degradation.

The Carlini–Wagner attack was evaluated separately because it minimizes perturbation magnitude rather than operating within a predefined perturbation bound. Robustness was summarized using ASR and the per-element RMS magnitudes of successful perturbations. More robust models were expected to exhibit lower ASRs and to require larger per-element RMS perturbations among successful attacks, whereas less robust models were expected to exhibit higher ASRs at smaller RMS distortions. ASR and RMS distortion were interpreted jointly because successful-attack distortion alone may reflect only the subset of examples for which the attack succeeded.

Black-box transferability was evaluated for medical models using PGD adversarial examples generated with perturbation bound *ɛ* =0.016. Each architecture was treated in turn as the white-box source model, and the resulting adversarial examples were evaluated against each of the other four target architectures.

We also visually inspected adversarial images from the weakest PGD adversarial test sets that reduced performance below 0.50 AUC in order to further understand the level of subtlety with which perturbations could significantly alter performance for each model.

### Adversarial Training

For adversarial training, the best checkpoint from each conventionally trained model, obtained using the procedure described above, was used to initialize 10 additional epochs of adversarial fine-tuning. During each training batch, PGD examples were generated online against the current model with an *ɛ*=0.016 perturbation bound and seven optimization steps with a step size of 0.004. PGD generation maximized classification cross-entropy for all architectures, while parameter updates used each model’s original architecture-specific training objective. Training batches consisted entirely of adversarial examples. Adversarially trained models were subsequently evaluated over the same PGD perturbation sweep as the conventionally trained models.

### Interpretability and Latent Space Analysis

To understand differences in model performance under adversarial perturbations and assess the characteristics of the most robust models, we analyzed each model’s feature space on original and adversarial inputs (Figure 1E). We used latent space and Gradient-weighted Class Activation Mapping (Grad-CAM) visualizations for this analysis.^58^ For all interpretability experiments, adversarial inputs were generated using a medium-strength PGD perturbation (*ɛ* = 0.016).

Latent space visualizations were used to assess how each model’s encoding of features altered under perturbation. For this analysis, activation feature vectors of all models were reduced to two dimensions using t-Distributed Stochastic Neighbor Embedding (t-SNE). Each reduced dimension was then normalized between 0 and 1. To ensure consistency between the original and adversarial embeddings, t-SNE embeddings for the original and adversarial latent spaces were calculated concurrently. To quantify the difference between the original and adversarial latent spaces for a particular model, cosine similarity was calculated between each image’s clean and adversarial feature vectors:

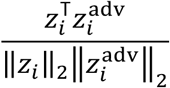

where zᵢ and zᵢᵃᵈᵛ denote the clean and adversarial feature vectors for image i, respectively. Similarities were averaged across the test set for each model and dataset. Higher cosine similarity scores would indicate greater preservation of the model’s internal representation under adversarial perturbation. Cosine similarity was calculated in the original, non-reduced feature space.

Grad-CAM visualizations were generated to further understand each model’s region of focus before and after adversarial perturbations. For ResNet models, Grad-CAMs were computed using the outputs of the final residual block. For the MedViT model, Grad-CAMs were applied to the grouped convolutional layer within the last Multi-Head Convolutional Attention block. For CapsNet models, the output of the convolutional layer was used. The target layers were selected to capture high-level semantic information while maintaining spatial resolution suitable for interpretability. To quantify a model’s ability to maintain its region of focus under perturbation, the average Intersection over Union (IoU) of the top 20% of activated pixels between each test set image and its perturbed counterpart was used. Models that lose focus under perturbation are expected to exhibit smaller average IoUs between their original and adversarial Grad-CAMs.

## Code Availability

Code for all experiments can be found at https://github.com/Aneja-Lab-Yale/Aneja-Lab-Public-Adversarial-CapsNet. All experiments were conducted on an Amazon EC2 g4dn.xlarge instance equipped with an NVIDIA Tesla T4 GPU (16 GB VRAM) and 4 vCPUs. All models were implemented in PyTorch 2.3.1 using Python 3.10.

## Data Availability

All MedMNIST datasets including PneumoniaMNIST, BreastMNIST, NoduleMNIST3D and BloodMNIST are publicly available for download at https://medmnist.com/. The original data sources for these datasets can be found at the following links: [PneumoniaMNIST](https://data.mendeley.com/datasets/rscbjbr9sj/3), [BreastMNIST](https://www.sciencedirect.com/science/article/pii/S2352340919312181?via%3Dihub), [NoduleMNIST3D](https://aapm.onlinelibrary.wiley.com/doi/10.1118/1.3528204) and [BloodMNIST](https://www.sciencedirect.com/science/article/pii/S2352340920303681?via%3Dihub). MNIST is publicly available for download from PyTorch at https://docs.pytorch.org/vision/main/generated/torchvision.datasets.MNIST.html.

## Results

### Adversarial Robustness of Capsule Networks

When compared to CNN and ViT models, CapsNet models were more robust to adversarial perturbations across all medical and natural image datasets (Figure 3). All models achieved similar performance at baseline (Supplementary Table 2). Against PGD perturbations, both BP-CapsNet and DR-CapsNet showed a slower decline in accuracy and AUC as the perturbation strength ε increased (Figure 3). In contrast, the ResNet and ViT models experienced sharp drops in performance, indicating limited adversarial robustness. For instance, at a moderate PGD perturbation of *ɛ*=0.016, BP-CapsNet and DR-CapsNet achieved AUCs ranging from 0.759–0.950 and from 0.728–0.915, respectively, across the four medical datasets. Within each dataset, the conventional architectures exhibited lower AUCs. ResNet-18 ranged from 0.287–0.823, ResNet-50 from 0.319–0.760, and MedViT from 0.247–0.754 (Table 2).

**Fig. 3.**
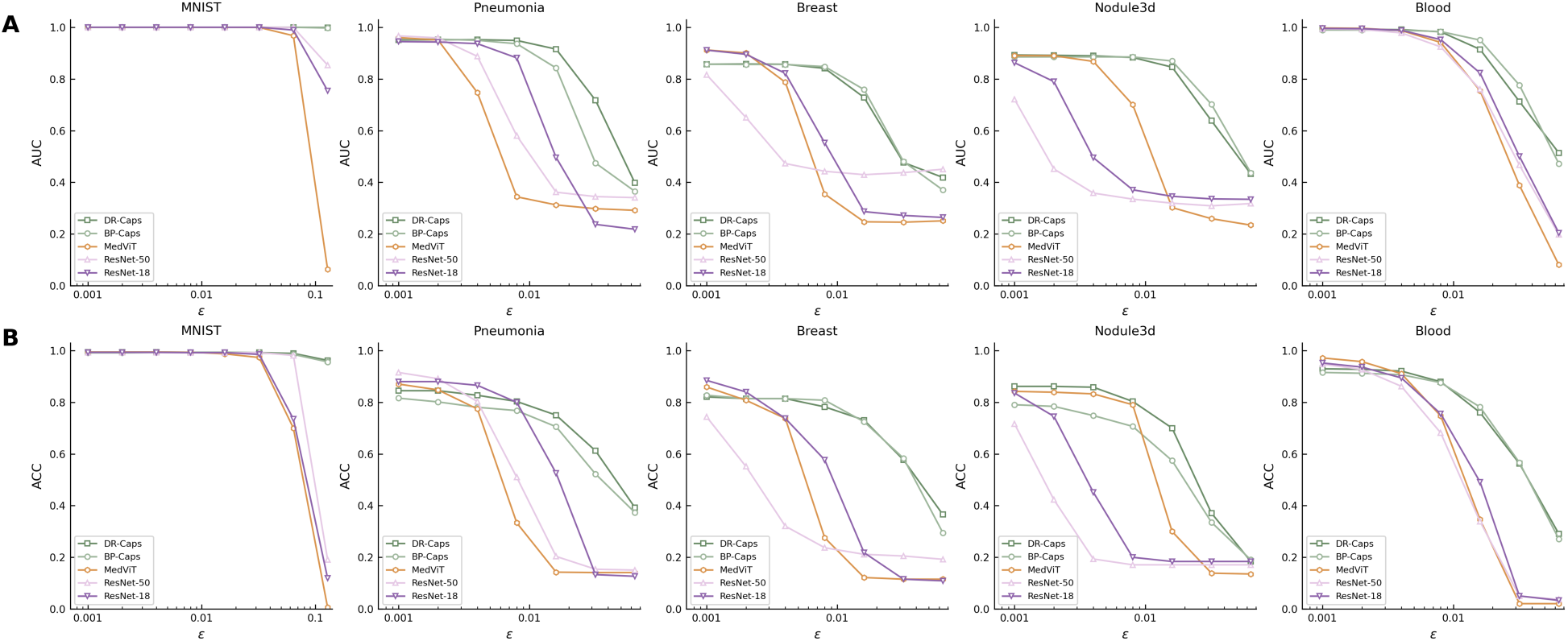
Model performance on projected gradient descent (PGD) adversarial images. **A.** Area under the receiver operating characteristic curve (AUC) scores for ResNet-18, ResNet-50, MedViT, DR-CapsNet, and BP-CapsNet models across natural and medical imaging datasets under PGD adversarial perturbations of increasing strength (ε). **B.** Accuracy (ACC) scores for ResNet-18, ResNet-50, MedViT, DR-CapsNet, and BP-CapsNet models across natural and medical imaging datasets under PGD adversarial perturbations of increasing strength (ε). Both AUC and accuracy plots display perturbation strengths up to ε = 0.128, with individual plots truncated once all models fall below 0.50 accuracy. At a moderate perturbation strength (ε = 0.016), BP-CapsNet and DR-CapsNet achieved AUC scores ranging from 0.759–0.950 and 0.728–0.915, respectively, across the four medical datasets. ResNet-18, ResNet-50, and MedViT performed degraded faster than CapsNets, achieving AUCs ranging from 0.287–0.823, 0.319–0.760, and 0.247–0.754, respectively, at the same moderate perturbation strength.

**Table 2.** Adversarial sensitivity results on projected gradient descent (PGD) adversarial images generated with varying perturbation bound *ɛ*.

| **Dataset** | ***ε*** | **ResNet-18** |  | **ResNet-50** |  | **MedViT** |  | **BP-CapsNet** |  | **DR-CapsNet** |  |
|---|---|---|---|---|---|---|---|---|---|---|---|
|  |  | AUC | ACC | AUC | ACC | AUC | ACC | AUC | ACC | AUC | ACC |
| **MNIST** | 0.000 | 1.000 | 0.993 | 1.000 | 0.993 | 1.000 | 0.994 | 1.000 | 0.991 | 1.000 | 0.994 |
|  | 0.001 | 1.000 | 0.993 | 1.000 | 0.993 | 1.000 | 0.994 | 1.000 | 0.991 | 1.000 | 0.994 |
|  | 0.002 | 1.000 | 0.993 | 1.000 | 0.993 | 1.000 | 0.994 | 1.000 | 0.991 | 1.000 | 0.994 |
|  | 0.004 | 1.000 | 0.993 | 1.000 | 0.992 | 1.000 | 0.994 | 1.000 | 0.992 | 1.000 | 0.994 |
|  | 0.008 | 1.000 | 0.992 | 1.000 | 0.991 | 1.000 | 0.993 | 1.000 | 0.992 | 1.000 | 0.993 |
|  | 0.016 | 1.000 | 0.992 | 1.000 | 0.991 | 1.000 | 0.987 | 1.000 | 0.992 | 1.000 | 0.993 |
|  | 0.032 | 1.000 | 0.985 | 1.000 | 0.992 | 0.999 | 0.973 | 1.000 | 0.990 | 1.000 | 0.992 |
|  | 0.064 | 0.989 | 0.735 | 0.999 | 0.981 | 0.967 | 0.700 | 1.000 | 0.985 | 1.000 | 0.989 |
|  | 0.128 | 0.754 | 0.119 | 0.853 | 0.191 | 0.063 | 0.006 | 0.997 | 0.956 | 0.999 | 0.962 |
| **Pneumonia MNIST** | 0.000 | 0.945 | 0.881 | 0.966 | 0.894 | 0.961 | 0.873 | 0.956 | 0.822 | 0.950 | 0.854 |
|  | 0.001 | 0.944 | 0.880 | 0.967 | 0.915 | 0.960 | 0.870 | 0.955 | 0.816 | 0.951 | 0.845 |
|  | 0.002 | 0.943 | 0.880 | 0.959 | 0.891 | 0.951 | 0.848 | 0.954 | 0.801 | 0.952 | 0.845 |
|  | 0.004 | 0.937 | 0.865 | 0.887 | 0.803 | 0.747 | 0.774 | 0.949 | 0.780 | 0.952 | 0.827 |
|  | 0.008 | 0.881 | 0.800 | 0.580 | 0.510 | 0.344 | 0.333 | 0.936 | 0.768 | 0.949 | 0.803 |
|  | 0.016 | 0.496 | 0.526 | 0.362 | 0.204 | 0.313 | 0.143 | 0.842 | 0.705 | 0.915 | 0.750 |
|  | 0.032 | 0.237 | 0.133 | 0.345 | 0.154 | 0.298 | 0.141 | 0.475 | 0.522 | 0.718 | 0.612 |
|  | 0.064 | 0.218 | 0.127 | 0.341 | 0.151 | 0.292 | 0.141 | 0.365 | 0.373 | 0.398 | 0.393 |
|  | 0.128 | 0.220 | 0.127 | 0.328 | 0.151 | 0.276 | 0.141 | 0.297 | 0.463 | 0.324 | 0.324 |
| **Breast MNIST** | 0.000 | 0.916 | 0.885 | 0.860 | 0.801 | 0.916 | 0.865 | 0.856 | 0.833 | 0.856 | 0.808 |
|  | 0.001 | 0.911 | 0.885 | 0.816 | 0.744 | 0.912 | 0.859 | 0.857 | 0.827 | 0.857 | 0.821 |
|  | 0.002 | 0.895 | 0.840 | 0.651 | 0.551 | 0.900 | 0.808 | 0.856 | 0.814 | 0.858 | 0.814 |
|  | 0.004 | 0.822 | 0.737 | 0.473 | 0.321 | 0.787 | 0.737 | 0.856 | 0.814 | 0.857 | 0.814 |
|  | 0.008 | 0.552 | 0.577 | 0.443 | 0.237 | 0.355 | 0.276 | 0.848 | 0.808 | 0.841 | 0.782 |
|  | 0.016 | 0.287 | 0.218 | 0.430 | 0.212 | 0.247 | 0.122 | 0.759 | 0.724 | 0.728 | 0.731 |
|  | 0.032 | 0.272 | 0.115 | 0.438 | 0.205 | 0.246 | 0.115 | 0.481 | 0.583 | 0.477 | 0.577 |
|  | 0.064 | 0.264 | 0.109 | 0.451 | 0.192 | 0.251 | 0.115 | 0.370 | 0.295 | 0.419 | 0.365 |
|  | 0.128 | 0.254 | 0.109 | 0.410 | 0.192 | 0.239 | 0.115 | 0.280 | 0.269 | 0.359 | 0.365 |
| **Nodule MNIST3D** | 0.000 | 0.861 | 0.819 | 0.868 | 0.832 | 0.888 | 0.848 | 0.886 | 0.806 | 0.894 | 0.861 |
|  | 0.001 | 0.863 | 0.835 | 0.721 | 0.716 | 0.890 | 0.842 | 0.885 | 0.790 | 0.893 | 0.861 |
|  | 0.002 | 0.790 | 0.745 | 0.452 | 0.423 | 0.890 | 0.839 | 0.885 | 0.784 | 0.891 | 0.861 |
|  | 0.004 | 0.496 | 0.452 | 0.358 | 0.194 | 0.868 | 0.832 | 0.885 | 0.748 | 0.889 | 0.858 |
|  | 0.008 | 0.371 | 0.200 | 0.335 | 0.171 | 0.700 | 0.790 | 0.885 | 0.706 | 0.883 | 0.803 |
|  | 0.016 | 0.346 | 0.184 | 0.319 | 0.171 | 0.302 | 0.300 | 0.869 | 0.574 | 0.846 | 0.700 |
|  | 0.032 | 0.336 | 0.184 | 0.309 | 0.171 | 0.259 | 0.139 | 0.702 | 0.335 | 0.638 | 0.371 |
|  | 0.064 | 0.334 | 0.184 | 0.318 | 0.171 | 0.234 | 0.135 | 0.437 | 0.190 | 0.432 | 0.184 |
|  | 0.128 | 0.352 | 0.184 | 0.287 | 0.171 | 0.226 | 0.135 | 0.288 | 0.171 | 0.293 | 0.161 |
| **Blood MNIST** | 0.000 | 0.996 | 0.958 | 0.995 | 0.953 | 0.998 | 0.975 | 0.989 | 0.917 | 0.993 | 0.931 |
|  | 0.001 | 0.995 | 0.952 | 0.994 | 0.947 | 0.997 | 0.971 | 0.989 | 0.915 | 0.992 | 0.929 |
|  | 0.002 | 0.994 | 0.936 | 0.992 | 0.927 | 0.996 | 0.957 | 0.989 | 0.912 | 0.992 | 0.927 |
|  | 0.004 | 0.988 | 0.894 | 0.978 | 0.862 | 0.987 | 0.911 | 0.988 | 0.906 | 0.991 | 0.920 |
|  | 0.008 | 0.952 | 0.755 | 0.924 | 0.682 | 0.941 | 0.747 | 0.983 | 0.876 | 0.982 | 0.879 |
|  | 0.016 | 0.823 | 0.491 | 0.760 | 0.338 | 0.754 | 0.347 | 0.950 | 0.781 | 0.914 | 0.760 |
|  | 0.032 | 0.500 | 0.051 | 0.467 | 0.050 | 0.389 | 0.021 | 0.776 | 0.567 | 0.713 | 0.562 |
|  | 0.064 | 0.204 | 0.033 | 0.198 | 0.037 | 0.081 | 0.021 | 0.472 | 0.271 | 0.514 | 0.291 |
|  | 0.128 | 0.091 | 0.029 | 0.125 | 0.031 | 0.030 | 0.018 | 0.306 | 0.039 | 0.367 | 0.056 |

Abbreviations: **PGD:** Projected Gradient Descent, **AUC:** Area Under the Receiver Operating Characteristic Curve, **ACC:** Classification Accuracy

CapsNet models also demonstrated the strongest robustness against FGSM adversarial images (Supplementary Figure 1). At a moderate FGSM perturbation (*ɛ*=0.016), the worst-case AUC observed across datasets was 0.767 for BP-CapsNet and 0.737 for DR-CapsNet. ResNet-18, ResNet-50, and MedViT had worst-case AUCs of 0.369, 0.362, and 0.262, respectively (Supplementary Table 3). An exception was observed for PneumoniaMNIST, where ResNet-50 outperformed BP-CapsNet but remained less robust than DR-CapsNet. FGSM generally produced less performance degradation than PGD, particularly for the ResNet and MedViT models. The capsule models remained comparatively robust under both attacks, with smaller and less consistent differences between their FGSM and PGD performance.

AutoAttack corroborated the robustness observed under PGD and FGSM across the four medical datasets (Supplementary Table 4). At *ɛ*=0.032, BP-CapsNet and DR-CapsNet achieved AUCs ranging from 0.604–0.843 and from 0.609–0.826, respectively. In comparison, AUCs ranged from 0.237–0.492, 0.304–0.419, and 0.248–0.353 for ResNet-18, ResNet-50, and MedViT models, respectively.

CW results at an initial attack constant of c=0.01 were consistent with greater adversarial robustness of the CapsNets (Supplementary Tables 5 and 6), which generally exhibited lower ASRs (1.1–80.5%) and higher median RMS distortions among successful attacks (0.0006–0.0106) than ResNet models (ASR: 98.4–100.0%; RMS: 0.0005–0.0064) and MedViT (ASR: 3.4–99.3%; RMS: 0.0003–0.0036). In a higher-constant convergence analysis (c=10), ASRs increased to 88.3–100.0% for BP-CapsNet and 80.3–100.0% for DR-CapsNet. These successful attacks were associated with greater median RMS distortions, ranging from 0.0086–0.0109 and 0.0078–0.0150, respectively. These findings indicate that CW ASR was sensitive to the attack constant, while successful attacks against the CapsNets generally required larger perturbations.

Black-box PGD perturbations generally transferred poorly between models, particularly across architecture families (Supplementary Table 7). Target-model AUCs remained between 0.735 and 0.994 across the medical datasets for the transfer attacks tested. Transfer was strongest between BP-CapsNet and DR-CapsNet, likely reflecting their architectural similarity.

Visual inspection of the adversarial images revealed that in order to reduce performance below a 0.50 AUC threshold, CapsNets required adversarial inputs that were more visibly altered, whereas the corresponding adversarial examples for the CNNs and ViT remained nearly indistinguishable from the originals (Figure 4).

**Fig. 4.**
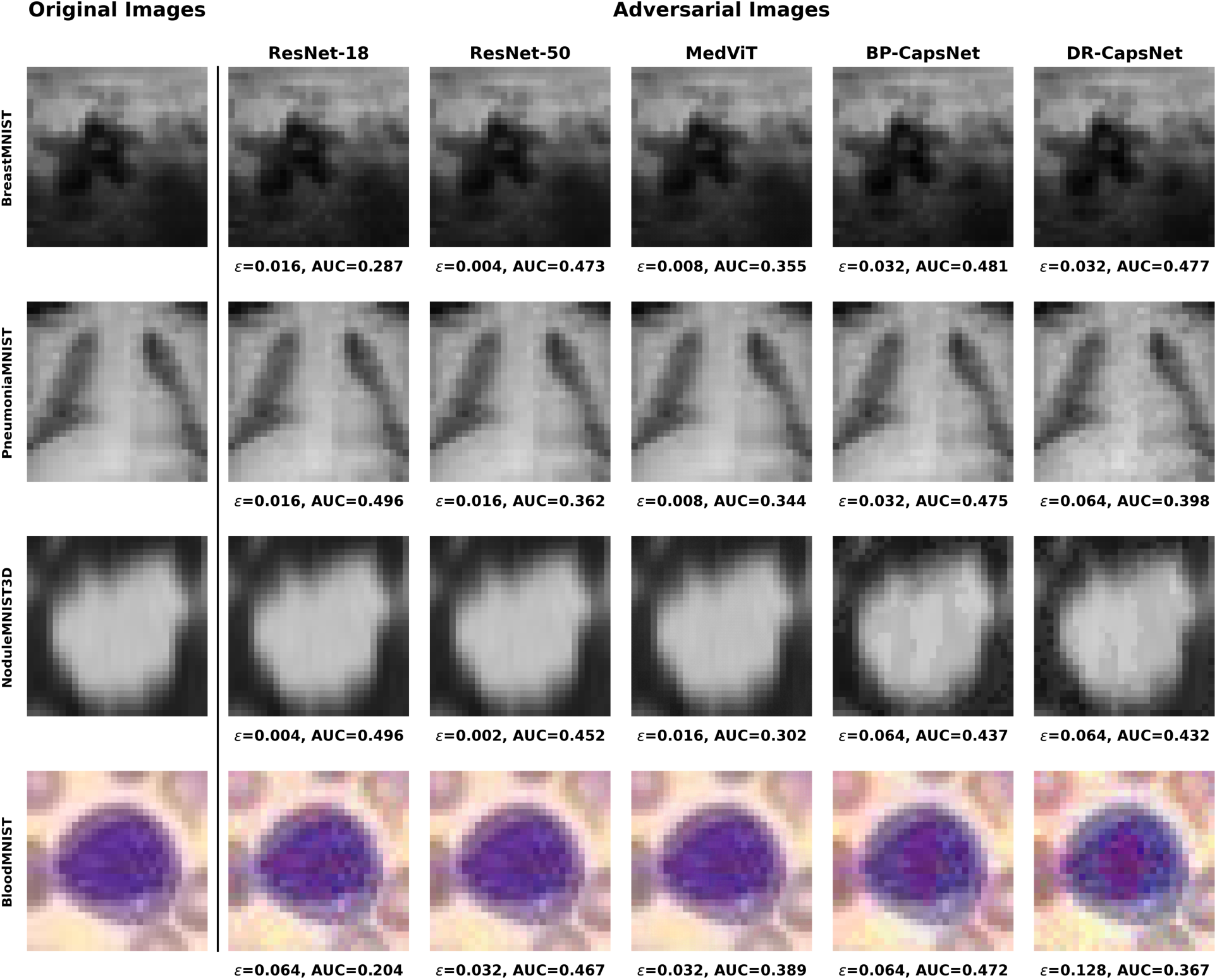
Examples of projected gradient descent (PGD) adversarial perturbations applied to achieve below an area under the receiver operating characteristic curve (AUC) of 0.50. Representative adversarial images according to the smallest tested perturbation size that produced under 0.50 AUC on the adversarial test set for each model across each medical dataset. Models were tested against PGD adversarial attacks of increasing strength following the *ɛ* values listed in Table 2. The observable perturbation required to achieve sub 0.50 AUC was greater for CapsNet models, especially for PneumoniaMNIST, NoduleMNIST3D, and BloodMNIST.

### Adversarial Training

PGD adversarial training improved robustness against PGD adversarial perturbations across most models (Supplementary Figure 2). MedViT benefited most from PGD adversarial training, with AUC gains of 0.232–0.645 at the training perturbation of ε=0.016. Across the broader perturbation sweep, the CapsNets generally maintained the strongest performance on BreastMNIST and PneumoniaMNIST, whereas MedViT demonstrated the greatest robustness on NoduleMNIST3D at perturbation strengths above the training epsilon. Overall, adversarial training narrowed the robustness gap between architectures, although CapsNet models remained the strongest performers on most datasets for which clear between-model differences persisted.

### Interpretability Experiments

#### Latent Space Visualizations

Latent space visualizations showed that CapsNet models possessed the more stable features compared to CNN and ViT models. Specifically, both CapsNet models retained better clustering of classes in their embedding spaces under adversarial perturbations compared to the CNNs and ViT (Figure 5G-J). CapsNet embeddings shifted less under perturbation, which was evidenced by higher mean cosine similarities between clean and adversarial feature vectors.

**Fig. 5.**
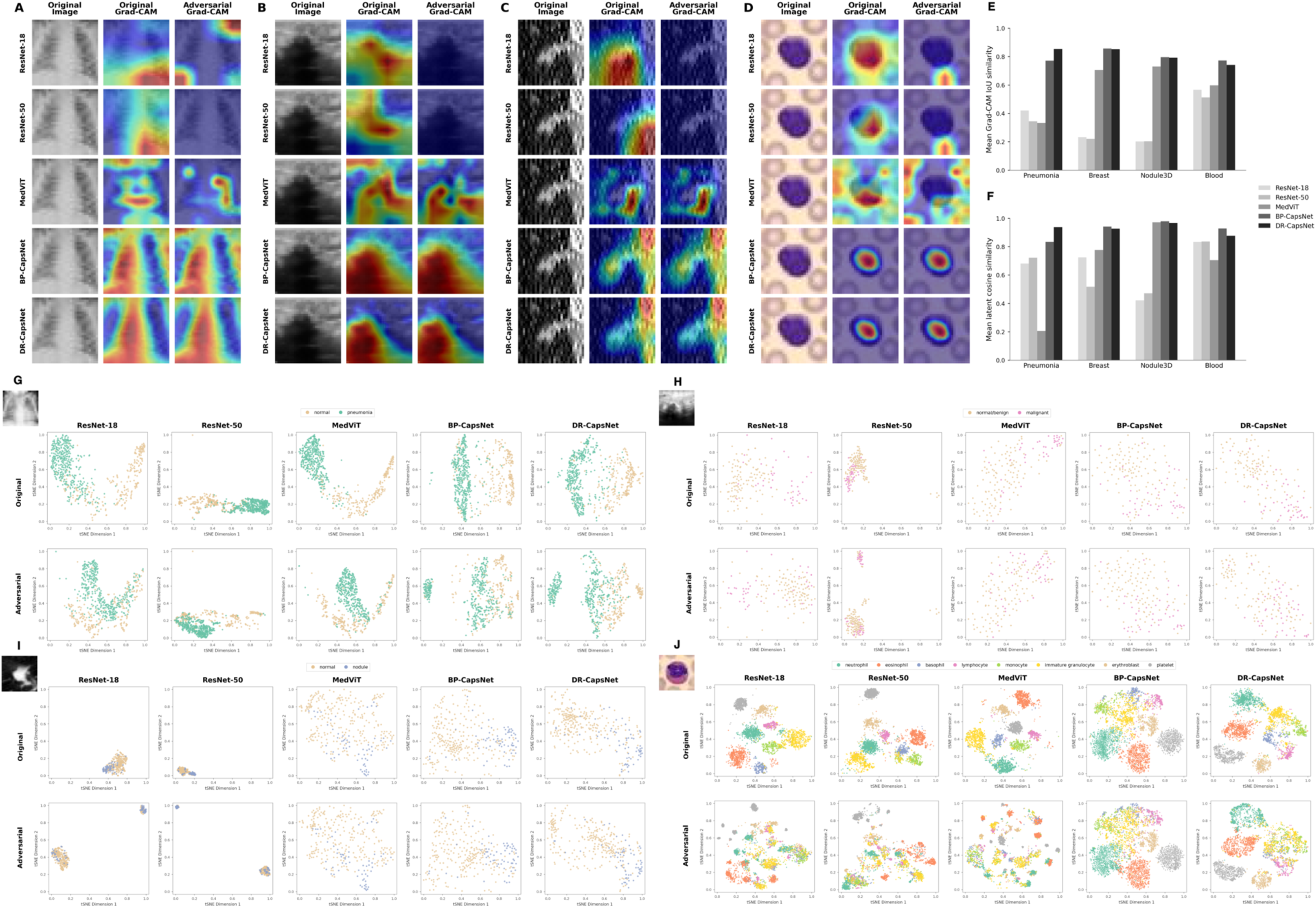
Interpretability analyses. **A-D.** Gradient-weighted Class Activation Mapping (Grad-CAMs) for all models on original and adversarial image pairs for (**A**) PneumoniaMNIST, (**B**) BreastMNIST, (**C**) NoduleMNIST3D, and (**D**) BloodMNIST datasets. Adversarial images were generated using projected gradient descent (PGD) with a perturbation bound of *ɛ* = 0.016. CapsNet Grad-CAMs remained consistent after adversarial perturbations, whereas Grad-CAMs for other models tended to shift in focus. **E.** Mean similarity between original Grad-CAMs and their adversarial counterparts for each model across each medical domain dataset. Similarity was measured as the intersection over union (IoU) between the top 20% of pixels for an original and adversarial Grad-CAM pair. CapsNet models exhibited the highest mean similarity between original and adversarial Grad-CAMs, demonstrating stability in their regions of focus. **F.** Cosine similarity between original examples and their adversarial counterparts in the feature space for each model across each medical domain dataset. CapsNet models exhibited the largest cosine similarities, indicating stable feature representations. **G-J.** First two components of the t-Distributed Stochastic Neighbor Embedding (t-SNE) projection of each model’s latent space on the original and adversarial test sets across (**G**) PneumoniaMNIST, (**H**) BreastMNIST, (**I**) NoduleMNIST3D, and (**J**) BloodMNIST datasets. CapsNet models retained latent space structure more effectively than ResNet and MedViT models following adversarial perturbations.

BP-CapsNet and DR-CapsNet mean cosine similarities ranged from 0.835–0.979 and 0.877–0.967, respectively, across the four medical datasets, compared with 0.421–0.834 for ResNet-18, 0.472–0.837 for ResNet-50, and 0.207–0.972 for MedViT (Figure 5F). Both CapsNet models exhibited higher cosine similarity than the ResNet models on every dataset; MedViT showed comparable representation stability on NoduleMNIST3D but substantially lower similarity on the other medical datasets.

#### Grad-CAM

Grad-CAM visualizations demonstrated that CapsNet models maintained more consistent focus on relevant image sections compared to CNN and ViT models. CapsNet models achieved higher mean Grad-CAM similarity scores between original and adversarial examples compared to CNN and ViT models, indicating improved preservation of model attention (Figure 5E). Across the medical imaging datasets, BP-CapsNet and DR-CapsNet similarity values ranged from 0.771–0.856 and 0.740–0.853, respectively, while ResNet-18, ResNet-50, and MedViT models produced mean similarity values in the ranges of 0.202–0.565, 0.202–0.512, and 0.332–0.729. Visually, CNN and ViT models exhibited substantial changes in their attention maps after perturbation, with attention moving away from meaningful image regions. In contrast, the Grad-CAM heatmaps of BP-CapsNet and DR-CapsNet models remained more stable after adversarial perturbations (Figure 5A-D, Supplementary Figure 3).

## Discussion

DL has had a profound impact on medical diagnostics, and its use is only expected to increase.^10,11^ As DL models become more mature safe integration into clinical workflows depends not only on high baseline performance but also on the ability of models to maintain stable and reliable predictions under real-world variability. Although adversarial perturbations are algorithmically constructed, they serve as a useful stress test for identifying model vulnerabilities that may overlap with clinically relevant sources of variation, including image noise, acquisition artifacts, and scanner heterogeneity. From a clinical perspective, models that are highly sensitive to small input perturbations may also be vulnerable to subtle but unavoidable differences in real-world imaging data.^22–24^ In this study, we observed that CapsNets consistently exhibited greater robustness to adversarial images over CNNs and ViTs across multiple medical imaging tasks. Importantly, these performance differences were observed across a range of black- and white-box adversarial attacks, perturbation strengths, and imaging modalities, suggesting that architectural design of a DL model may play a fundamental role in determining model stability.

A key contribution of this work is the observation that improved adversarial robustness in CapsNets is accompanied by increased stability of internal feature representations. Latent space analyses demonstrated that CapsNet embeddings shift less under perturbation, while Grad-CAM visualizations showed greater consistency in regions of model attention compared to CNN and ViT models. Together, these results suggest that CapsNets maintain more invariant representations in the presence of input perturbations. This representational stability provides a plausible mechanistic explanation for their improved robustness and may reflect the ability of capsule-based architectures to encode spatial relationships and hierarchical features in a manner that is less sensitive to small, targeted perturbations. These findings move beyond performance benchmarking and offer insight into how architectural properties influence model behavior under stress.

Previous studies on non-medical images have similarly suggested CapsNets to be more robust to adversarial inputs. On adversarial images generated by basic iterative method and FGSM, matrix CapsNets have been shown to exhibit stronger performance compared to a simple three-layer CNN model. Standard CapsNet models have similarly been found to demonstrate greater robustness to PGD and Carlini-Wagner adversarial inputs than deep CNNs.^44,46^ Our work builds upon these findings, focusing on CapsNet stability within the medical imaging domain, where models have shown unique sensitivity to adversarial perturbations.^27^ We demonstrated that CapsNets exhibit superior performance on adversarial images not only relative to simple CNNs but also to state-of-the-art CNN and ViT models. We also performed Grad-CAM and latent space interpretability experiments and illustrated that CapsNet features remain more consistent than CNNs and ViTs after perturbation, suggesting a potential explanation for their increased stability.

Some prior work has shown that CapsNets have underperformed against adversarial images. In Michels et al., a standard CapsNet outperformed a CNN on the MNIST dataset across all perturbation methods tested, consistent with our findings.^45^ CapsNets, however, did not demonstrate stronger adversarial performance for the other datasets evaluated in their study (Fashion-MNIST, SVHN, CIFAR-10). For these datasets, additional convolutional components were introduced into the CapsNet architecture to maintain high baseline fidelity. These hybrid convolutional CapsNets exhibited decreased adversarial resilience. Together with our findings, we conclude that standard CapsNets demonstrate improved performance on adversarial images compared to CNNs, but the introduction of additional convolutional components may negate the benefits conferred by capsule-based representations.

ViT–based models have previously been described to exhibit increased resistance to adversarial perturbations compared to CNNs.^55,59^ However, this behavior was not consistently observed in our study. Although MedViT has been reported to provide modest improvements in robustness over CNNs under FGSM and PGD attacks, prior evaluations focused on a single perturbation bound ε and a smaller MedViT variant.^55^ In contrast, we assessed robustness across a range of ε values to characterize performance trends under varying perturbation strengths, which may partly explain the observed discrepancies. While ViTs have demonstrated superior stability in other domains, this advantage did not consistently translate to the medical imaging setting examined in our study.^59,60^ One possible explanation is that the MedViT’s hybrid architecture, while advantageous for medical image classification performance, may attenuate gains in robustness that have been attributed to purely transformer-based designs.

Adversarial training with PGD fine-tuning generally improved performance across model families, with MedViT exhibiting the largest gains and thereby narrowing the robustness gap between architectures. The substantial improvement observed for MedViT warrants further investigation into whether transformer-based architectures are particularly responsive to adversarial training. Nevertheless, adversarially trained CapsNets generally outperformed ResNets and MedViT on most datasets for which clear differences persisted, suggesting that their architectural robustness provides a favorable starting point that remains beneficial after adversarial training. Future work could explore CapsNet-specific adversarial-training strategies that account for capsule routing and representation structure. Because both adversarial training and subsequent evaluation used PGD attacks, future studies should also determine whether these improvements generalize to non-PGD attack types.

Our study has several limitations. Prior work has suggested that capsule-routing mechanisms may produce gradient masking, potentially reducing the effectiveness of certain gradient-based attacks.^44,45^ We sought to mitigate this concern by including gradient-free black-box attacks in our evaluation, including black-box transfer attacks and an AutoAttack ensemble which incorporated Square Attacks. More broadly, our evaluation was limited to FGSM, PGD, CW, AutoAttack, and transfer attacks, and the observed robustness may not generalize to all types of adversarial attacks. Nevertheless, these methods are among the most widely used adversarial perturbation methods and serve as standard benchmarks for evaluating adversarial robustness and model stability, including in medical imaging contexts.^56,57^ Future work should investigate additional adversarial perturbation methods that could provide insight beyond what is captured in our study. This study also only considered DL models for image classification tasks and did not investigate other well-known imaging tasks that leverage DL, namely, segmentation and image reconstruction. The benefits in robustness of medical CapsNets for image classification may not generalize to these other tasks, and further studies to evaluate adversarial performance for medical segmentation and image reconstruction are needed. Lastly, our study was limited to standardized, low-resolution benchmark datasets. Evaluation using full-resolution medical images and independent external datasets is needed before drawing definitive conclusions about the generalizability and robustness conferred by model architecture.

In conclusion, we demonstrate that CapsNet architectures exhibit improved robustness to adversarial perturbations and maintain more stable internal representations compared to CNN- and ViT-based models across multiple medical imaging tasks. These findings suggest that architectural design plays a critical role in determining model stability and highlight CapsNets as a promising alternative for developing more reliable medical AI systems. As the field moves toward real-world deployment of DL models in clinical practice, incorporating robustness considerations into model design and evaluation will be essential. Our results support the use of architecture-driven approaches as part of a broader strategy to enhance the safety and clinical trustworthiness of medical AI.

---

**Suppl. Table 1.** Model characteristics.

| **Model** | **Model Size<br>(10<sup>6</sup> parameters)** |  | **Inference Latency<br>(ms/image)** |  |  |  |
|---|---|---|---|---|---|---|
|  | 2D | 3D | PneumoniaMNIST | BreastMNIST | NoduleMNIST3D | BloodMNIST |
| ResNet-18 | 11.17 | 11.18 | 0.26 | 0.26 | 0.27 | 0.26 |
| ResNet-50 | 23.50 | 23.52 | 0.81 | 0.81 | 0.82 | 0.82 |
| MedViT | 31.14 | 31.15 | 10.10 | 10.09 | 10.22 | 10.10 |
| BP-CapsNet | 6.97 | 29.23 | 0.30 | 0.30 | 0.34 | 0.50 |
| DR-CapsNet | 6.97 | 29.23 | 0.28 | 0.28 | 0.31 | 0.34 |

Note: Inference latency times represent median classification-only inference latency in milliseconds per image. Measurements were obtained on an NVIDIA Tesla T4 over 200 iterations with a batch size of 16, following 50 warm-up iterations and exclude data loading, and preprocessing.

**Suppl. Table 2.**
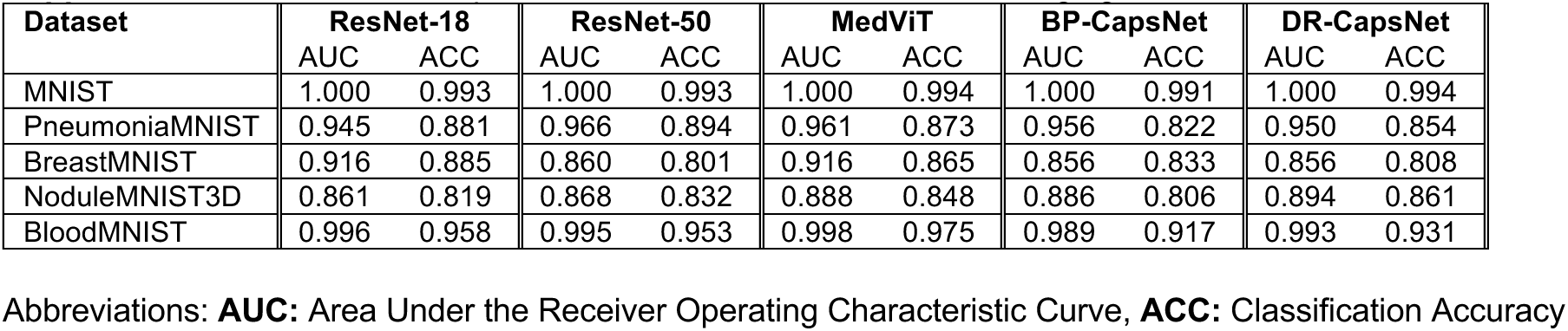
Baseline model performance on natural and medical imaging datasets.

| **Dataset** | **ResNet-18** |  | **ResNet-50** |  | **MedViT** |  | **BP-CapsNet** |  | **DR-CapsNet** |  |
|---|---|---|---|---|---|---|---|---|---|---|
|  | AUC | ACC | AUC | ACC | AUC | ACC | AUC | ACC | AUC | ACC |
| MNIST | 1.000 | 0.993 | 1.000 | 0.993 | 1.000 | 0.994 | 1.000 | 0.991 | 1.000 | 0.994 |
| PneumoniaMNIST | 0.945 | 0.881 | 0.966 | 0.894 | 0.961 | 0.873 | 0.956 | 0.822 | 0.950 | 0.854 |
| BreastMNIST | 0.916 | 0.885 | 0.860 | 0.801 | 0.916 | 0.865 | 0.856 | 0.833 | 0.856 | 0.808 |
| NoduleMNIST3D | 0.861 | 0.819 | 0.868 | 0.832 | 0.888 | 0.848 | 0.886 | 0.806 | 0.894 | 0.861 |
| BloodMNIST | 0.996 | 0.958 | 0.995 | 0.953 | 0.998 | 0.975 | 0.989 | 0.917 | 0.993 | 0.931 |

Abbreviations: **AUC:** Area Under the Receiver Operating Characteristic Curve, **ACC:** Classification Accuracy

**Suppl. Fig. 1.**
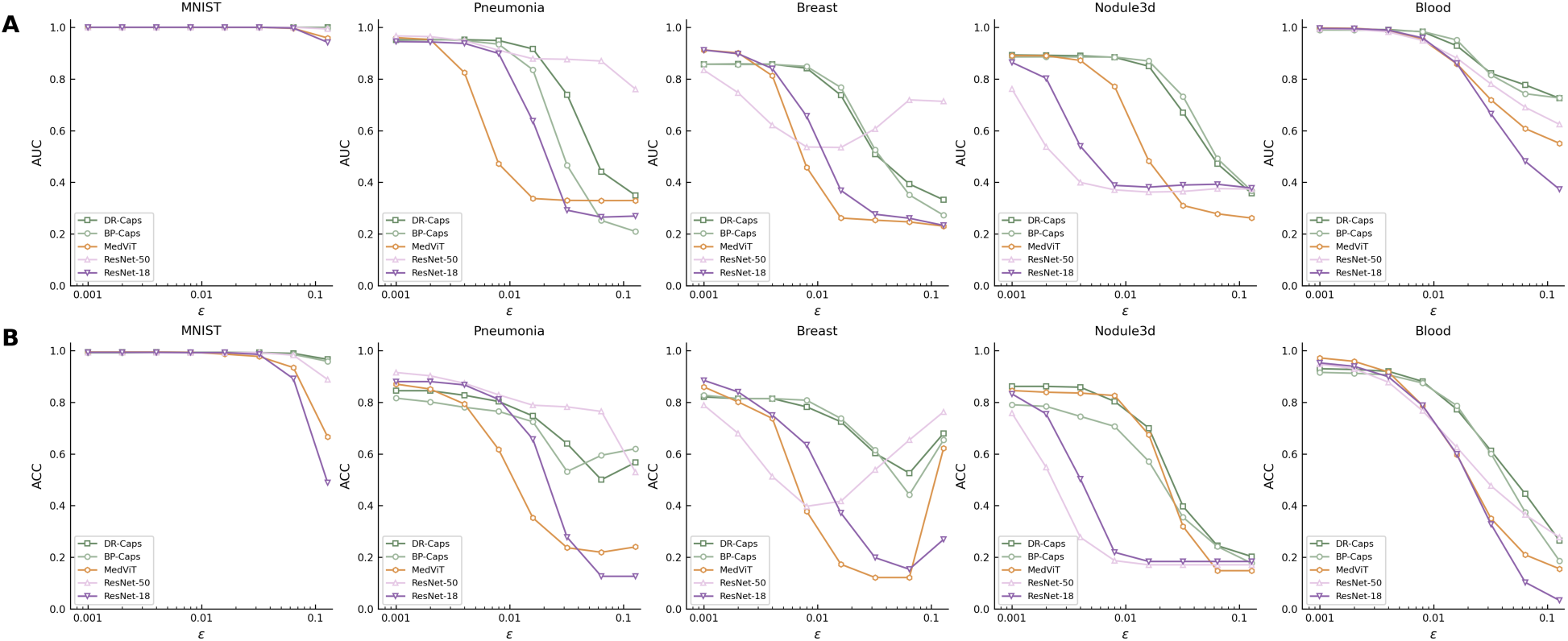
Model performance on fast gradient sign method (FGSM) adversarial images. **A.** Area under the receiver operating characteristic curve (AUC) scores for ResNet-18, ResNet-50, MedViT, DR-CapsNet, and BP-CapsNet models across natural and medical imaging datasets under FGSM adversarial perturbations of increasing strength (ε). **B.** Accuracy (ACC) scores for ResNet-18, ResNet-50, MedViT, DR-CapsNet, and BP-CapsNet models across natural and medical imaging datasets under FGSM adversarial perturbations of increasing strength (ε). Performance degraded faster for the ResNet and MedViT models compared with the CapsNets. At a moderate-strength perturbation (ε = 0.016), BP-CapsNet and DR-CapsNet achieved AUC scores ranging from 0.767–0.951 and 0.737–0.928, respectively, across the four medical datasets. ResNet-18, ResNet-50, and MedViT achieved AUCs ranging from 0.369–0.860, 0.362–0.880, and 0.262–0.858, respectively.

**Suppl. Table 3.** Adversarial sensitivity results on fast gradient sign method (FGSM) adversarial images with varying perturbation bound *ɛ*.

| **Dataset** | ***ε*** | **ResNet-18** |  | **ResNet-50** |  | **MedViT** |  | **BP-CapsNet** |  | **DR-CapsNet** |  |
|---|---|---|---|---|---|---|---|---|---|---|---|
|  |  | AUC | ACC | AUC | ACC | AUC | ACC | AUC | ACC | AUC | ACC |
| MNIST | 0.000 | 1.000 | 0.993 | 1.000 | 0.993 | 1.000 | 0.994 | 1.000 | 0.991 | 1.000 | 0.994 |
|  | 0.001 | 1.000 | 0.993 | 1.000 | 0.993 | 1.000 | 0.994 | 1.000 | 0.991 | 1.000 | 0.994 |
|  | 0.002 | 1.000 | 0.993 | 1.000 | 0.993 | 1.000 | 0.994 | 1.000 | 0.991 | 1.000 | 0.994 |
|  | 0.004 | 1.000 | 0.993 | 1.000 | 0.992 | 1.000 | 0.994 | 1.000 | 0.992 | 1.000 | 0.994 |
|  | 0.008 | 1.000 | 0.992 | 1.000 | 0.991 | 1.000 | 0.993 | 1.000 | 0.992 | 1.000 | 0.993 |
|  | 0.016 | 1.000 | 0.992 | 1.000 | 0.991 | 1.000 | 0.986 | 1.000 | 0.992 | 1.000 | 0.993 |
|  | 0.032 | 1.000 | 0.985 | 1.000 | 0.992 | 1.000 | 0.977 | 1.000 | 0.990 | 1.000 | 0.992 |
|  | 0.064 | 0.997 | 0.891 | 1.000 | 0.982 | 0.996 | 0.934 | 1.000 | 0.985 | 1.000 | 0.989 |
|  | 0.128 | 0.942 | 0.489 | 0.993 | 0.888 | 0.958 | 0.666 | 0.997 | 0.958 | 0.999 | 0.966 |
| Pneumonia MNIST | 0.000 | 0.945 | 0.881 | 0.966 | 0.894 | 0.961 | 0.873 | 0.956 | 0.822 | 0.950 | 0.854 |
|  | 0.001 | 0.944 | 0.880 | 0.967 | 0.915 | 0.960 | 0.870 | 0.955 | 0.816 | 0.951 | 0.845 |
|  | 0.002 | 0.943 | 0.880 | 0.964 | 0.902 | 0.953 | 0.851 | 0.954 | 0.801 | 0.952 | 0.845 |
|  | 0.004 | 0.938 | 0.867 | 0.948 | 0.873 | 0.824 | 0.793 | 0.949 | 0.780 | 0.952 | 0.827 |
|  | 0.008 | 0.899 | 0.811 | 0.911 | 0.829 | 0.472 | 0.617 | 0.934 | 0.764 | 0.949 | 0.803 |
|  | 0.016 | 0.637 | 0.657 | 0.878 | 0.789 | 0.337 | 0.353 | 0.835 | 0.724 | 0.916 | 0.747 |
|  | 0.032 | 0.292 | 0.277 | 0.876 | 0.782 | 0.330 | 0.237 | 0.466 | 0.532 | 0.739 | 0.639 |
|  | 0.064 | 0.265 | 0.127 | 0.869 | 0.764 | 0.329 | 0.220 | 0.253 | 0.595 | 0.441 | 0.500 |
|  | 0.128 | 0.270 | 0.127 | 0.760 | 0.529 | 0.330 | 0.240 | 0.210 | 0.620 | 0.349 | 0.567 |
| Breast MNIST | 0.000 | 0.916 | 0.885 | 0.860 | 0.801 | 0.916 | 0.865 | 0.856 | 0.833 | 0.856 | 0.808 |
|  | 0.001 | 0.912 | 0.885 | 0.834 | 0.788 | 0.912 | 0.859 | 0.857 | 0.827 | 0.857 | 0.821 |
|  | 0.002 | 0.897 | 0.840 | 0.746 | 0.679 | 0.901 | 0.801 | 0.856 | 0.814 | 0.858 | 0.814 |
|  | 0.004 | 0.840 | 0.750 | 0.621 | 0.513 | 0.812 | 0.737 | 0.856 | 0.814 | 0.857 | 0.814 |
|  | 0.008 | 0.656 | 0.635 | 0.537 | 0.397 | 0.458 | 0.378 | 0.849 | 0.808 | 0.842 | 0.782 |
|  | 0.016 | 0.369 | 0.372 | 0.535 | 0.417 | 0.262 | 0.173 | 0.767 | 0.737 | 0.737 | 0.724 |
|  | 0.032 | 0.277 | 0.199 | 0.607 | 0.538 | 0.254 | 0.122 | 0.526 | 0.615 | 0.509 | 0.603 |
|  | 0.064 | 0.261 | 0.154 | 0.719 | 0.654 | 0.247 | 0.122 | 0.351 | 0.442 | 0.393 | 0.526 |
|  | 0.128 | 0.234 | 0.269 | 0.713 | 0.763 | 0.231 | 0.622 | 0.273 | 0.654 | 0.332 | 0.679 |
| Nodule MNIST3D | 0.000 | 0.861 | 0.819 | 0.868 | 0.832 | 0.888 | 0.848 | 0.886 | 0.806 | 0.894 | 0.861 |
|  | 0.001 | 0.864 | 0.832 | 0.761 | 0.758 | 0.890 | 0.845 | 0.885 | 0.790 | 0.893 | 0.861 |
|  | 0.002 | 0.802 | 0.755 | 0.538 | 0.548 | 0.890 | 0.839 | 0.885 | 0.784 | 0.891 | 0.861 |
|  | 0.004 | 0.540 | 0.503 | 0.400 | 0.277 | 0.872 | 0.835 | 0.885 | 0.745 | 0.889 | 0.858 |
|  | 0.008 | 0.388 | 0.219 | 0.371 | 0.187 | 0.771 | 0.826 | 0.885 | 0.706 | 0.884 | 0.803 |
|  | 0.016 | 0.382 | 0.184 | 0.362 | 0.171 | 0.482 | 0.674 | 0.869 | 0.571 | 0.849 | 0.700 |
|  | 0.032 | 0.390 | 0.184 | 0.365 | 0.171 | 0.310 | 0.319 | 0.732 | 0.355 | 0.670 | 0.397 |
|  | 0.064 | 0.393 | 0.184 | 0.376 | 0.171 | 0.278 | 0.148 | 0.492 | 0.242 | 0.472 | 0.245 |
|  | 0.128 | 0.378 | 0.184 | 0.374 | 0.171 | 0.262 | 0.148 | 0.367 | 0.177 | 0.357 | 0.203 |
| Blood MNIST | 0.000 | 0.996 | 0.958 | 0.995 | 0.953 | 0.998 | 0.975 | 0.989 | 0.917 | 0.993 | 0.931 |
|  | 0.001 | 0.996 | 0.952 | 0.994 | 0.947 | 0.997 | 0.971 | 0.989 | 0.916 | 0.992 | 0.930 |
|  | 0.002 | 0.994 | 0.939 | 0.992 | 0.930 | 0.996 | 0.958 | 0.989 | 0.912 | 0.992 | 0.927 |
|  | 0.004 | 0.989 | 0.899 | 0.982 | 0.877 | 0.988 | 0.917 | 0.988 | 0.906 | 0.991 | 0.920 |
|  | 0.008 | 0.959 | 0.787 | 0.949 | 0.767 | 0.952 | 0.787 | 0.983 | 0.874 | 0.983 | 0.880 |
|  | 0.016 | 0.860 | 0.599 | 0.880 | 0.626 | 0.858 | 0.597 | 0.951 | 0.787 | 0.928 | 0.773 |
|  | 0.032 | 0.665 | 0.328 | 0.780 | 0.476 | 0.719 | 0.350 | 0.815 | 0.601 | 0.822 | 0.612 |
|  | 0.064 | 0.481 | 0.103 | 0.690 | 0.365 | 0.608 | 0.210 | 0.743 | 0.375 | 0.777 | 0.445 |
|  | 0.128 | 0.373 | 0.033 | 0.624 | 0.277 | 0.551 | 0.156 | 0.726 | 0.186 | 0.725 | 0.264 |

Abbreviations: **FGSM:** Fast Gradient Sign Method, **AUC:** Area Under the Receiver Operating Characteristic Curve, **ACC:** Classification Accuracy

**Suppl. Table 4.**
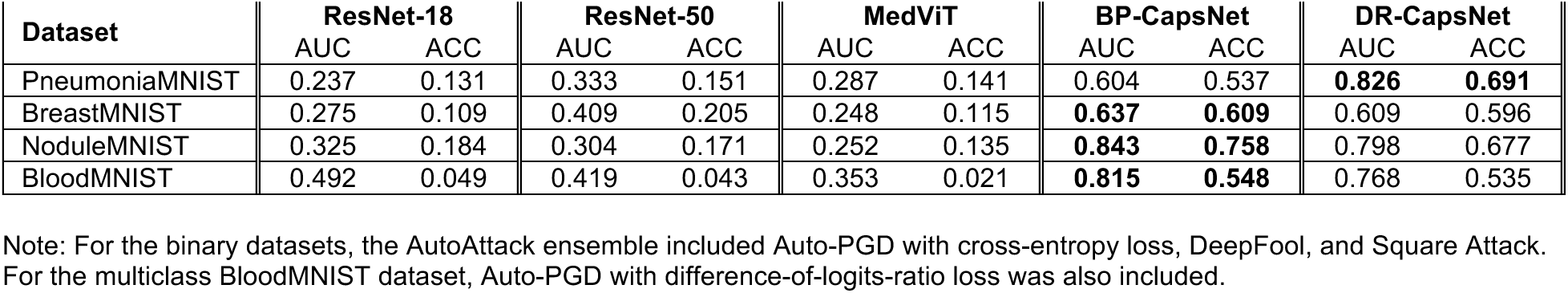
Adversarial sensitivity results on Auto Attack (*ɛ* = 0.032) adversarial images.

**Suppl. Table 5.** Adversarial sensitivity results on Carlini & Wagner adversarial images with initial trade-off constant c₀ = 0.01.

| **Model** | **PneumoniaMNIST** |  | **BreastMNIST** |  | **NoduleMNIST** |  | **BloodMNIST** |  |
|---|---|---|---|---|---|---|---|---|
|  | ASR (%) | Median RMS | ASR (%) | Median RMS | ASR (%) | Median RMS | ASR (%) | Median RMS |
| ResNet-18 | 99.82 | 0.0064 | 100.00 | 0.0031 | 98.42 | 0.0013 | 99.00 | 0.0048 |
| ResNet-50 | 100.00 | 0.0029 | 100.00 | 0.0008 | 99.61 | 0.0005 | 99.67 | 0.0038 |
| MedViT | 99.07 | 0.0016 | 99.28 | 0.0015 | 3.36 | 0.0003 | 98.33 | 0.0036 |
| BP-CapsNet | 70.30 | 0.0078 | 80.47 | 0.0082 | 1.13 | 0.0006 | 58.33 | 0.0054 |
| DR-CapsNet | 70.52 | 0.0106 | 74.80 | 0.0074 | 4.80 | 0.0014 | 54.00 | 0.0049 |

**Suppl. Table 6.**
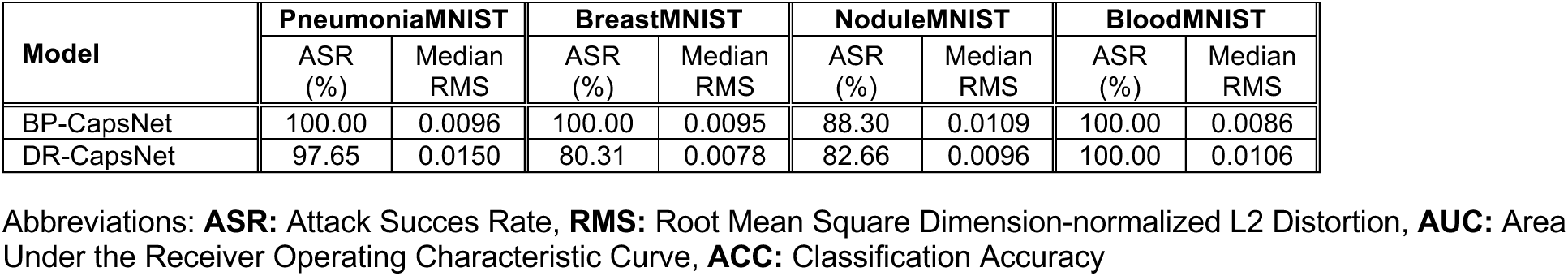
Additional adversarial sensitivity results on Carlini & Wagner adversarial images with initial trade-off constant c₀ = 10.

**Suppl. Table 7.**
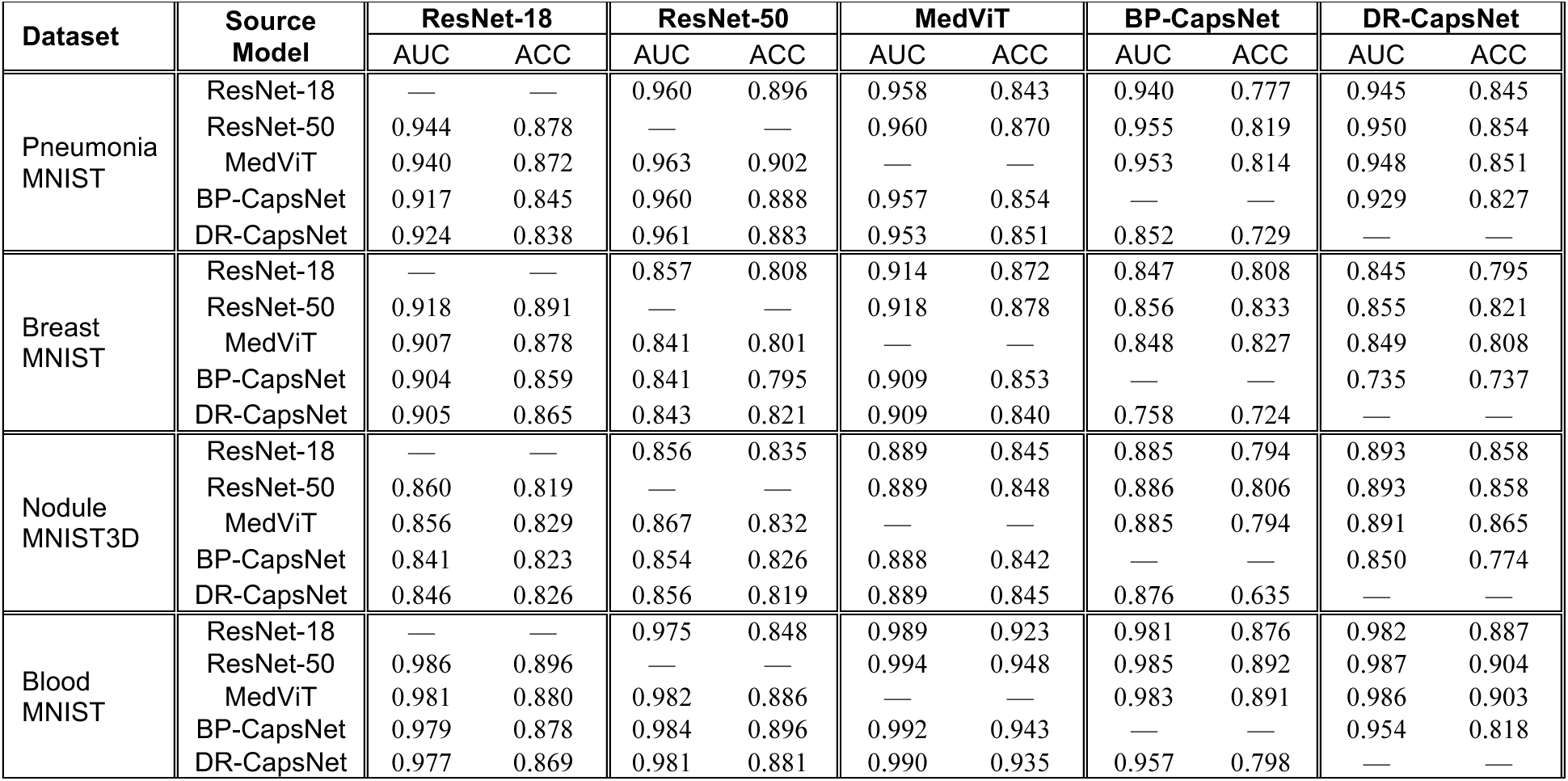
Adversarial sensitivity results on PGD (*ɛ* = 0.016) generated black box transfer attacks.

| **Dataset** | **Source Model** | **ResNet-18** |  | **ResNet-50** |  | **MedViT** |  | **BP-CapsNet** |  | **DR-CapsNet** |  |
|---|---|---|---|---|---|---|---|---|---|---|---|
|  |  | AUC | ACC | AUC | ACC | AUC | ACC | AUC | ACC | AUC | ACC |
| Pneumonia MNIST | ResNet-18 | — | — | 0.960 | 0.896 | 0.958 | 0.843 | 0.940 | 0.777 | 0.945 | 0.845 |
|  | ResNet-50 | 0.944 | 0.878 | — | — | 0.960 | 0.870 | 0.955 | 0.819 | 0.950 | 0.854 |
|  | MedViT | 0.940 | 0.872 | 0.963 | 0.902 | — | — | 0.953 | 0.814 | 0.948 | 0.851 |
|  | BP-CapsNet | 0.917 | 0.845 | 0.960 | 0.888 | 0.957 | 0.854 | — | — | 0.929 | 0.827 |
|  | DR-CapsNet | 0.924 | 0.838 | 0.961 | 0.883 | 0.953 | 0.851 | 0.852 | 0.729 | — | — |
| Breast MNIST | ResNet-18 | — | — | 0.857 | 0.808 | 0.914 | 0.872 | 0.847 | 0.808 | 0.845 | 0.795 |
|  | ResNet-50 | 0.918 | 0.891 | — | — | 0.918 | 0.878 | 0.856 | 0.833 | 0.855 | 0.821 |
|  | MedViT | 0.907 | 0.878 | 0.841 | 0.801 | — | — | 0.848 | 0.827 | 0.849 | 0.808 |
|  | BP-CapsNet | 0.904 | 0.859 | 0.841 | 0.795 | 0.909 | 0.853 | — | — | 0.735 | 0.737 |
|  | DR-CapsNet | 0.905 | 0.865 | 0.843 | 0.821 | 0.909 | 0.840 | 0.758 | 0.724 | — | — |
| Nodule MNIST3D | ResNet-18 | — | — | 0.856 | 0.835 | 0.889 | 0.845 | 0.885 | 0.794 | 0.893 | 0.858 |
|  | ResNet-50 | 0.860 | 0.819 | — | — | 0.889 | 0.848 | 0.886 | 0.806 | 0.893 | 0.858 |
|  | MedViT | 0.856 | 0.829 | 0.867 | 0.832 | — | — | 0.885 | 0.794 | 0.891 | 0.865 |
|  | BP-CapsNet | 0.841 | 0.823 | 0.854 | 0.826 | 0.888 | 0.842 | — | — | 0.850 | 0.774 |
|  | DR-CapsNet | 0.846 | 0.826 | 0.856 | 0.819 | 0.889 | 0.845 | 0.876 | 0.635 | — | — |
| Blood MNIST | ResNet-18 | — | — | 0.975 | 0.848 | 0.989 | 0.923 | 0.981 | 0.876 | 0.982 | 0.887 |
|  | ResNet-50 | 0.986 | 0.896 | — | — | 0.994 | 0.948 | 0.985 | 0.892 | 0.987 | 0.904 |
|  | MedViT | 0.981 | 0.880 | 0.982 | 0.886 | — | — | 0.983 | 0.891 | 0.986 | 0.903 |
|  | BP-CapsNet | 0.979 | 0.878 | 0.984 | 0.896 | 0.992 | 0.943 | — | — | 0.954 | 0.818 |
|  | DR-CapsNet | 0.977 | 0.869 | 0.981 | 0.881 | 0.990 | 0.935 | 0.957 | 0.798 | — | — |

**Suppl. Table 8.**
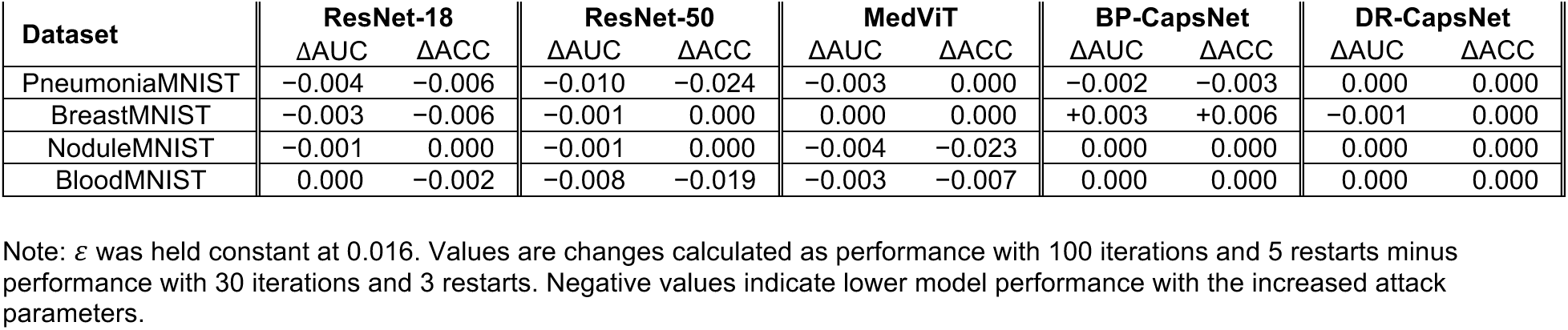
Change in model performance after increasing PGD attack iterations and restarts from 30 iterations and 3 restarts to 100 iterations and 5 restarts.

**Suppl. Fig. 2.**
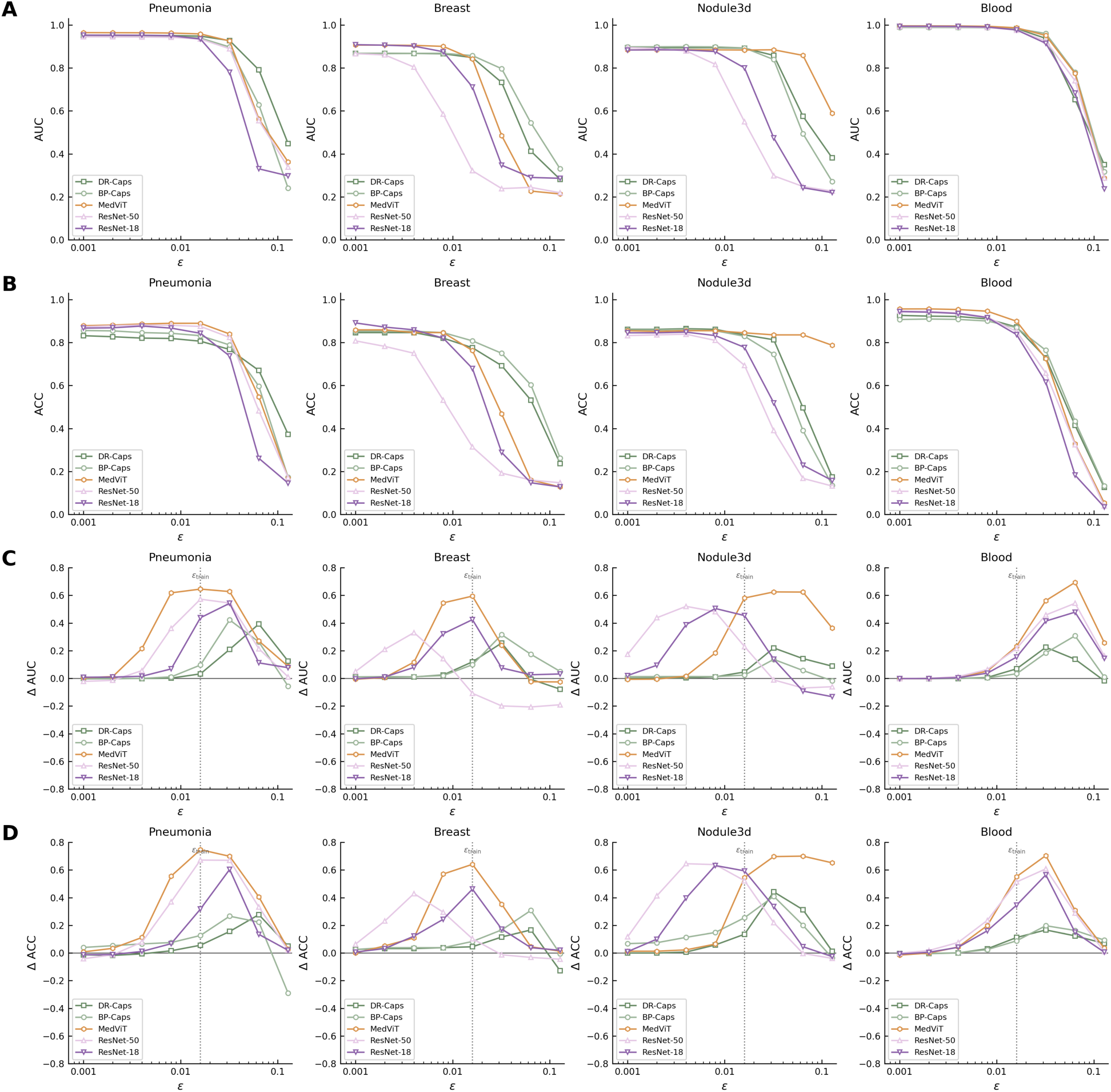
Adversarial training. **A-B.** Test-set AUC (**A**) and accuracy (**B**) of adversarially trained models under PGD attacks of increasing strength. **C-D.** Corresponding change in AUC (**C**) and accuracy (**D**) relative to standard training (adversarially trained minus standard), with the dotted line indicating the training budget, *ɛ*train = 0.016. During training, each mini-batch was replaced by adversarial examples generated using label-aware, untargeted 7-step PGD with one random start. Evaluation used 30-step PGD with three random restarts. Adversarial training hardened all models, although gains varied with perturbation strength, and generally produced the largest improvements for MedViT. Following adversarial training, DR-Caps and BP-Caps retained the strongest robustness on PneumoniaMNIST and BreastMNIST, whereas MedViT performed best on NoduleMNIST3D.

**Suppl. Fig. 3.**
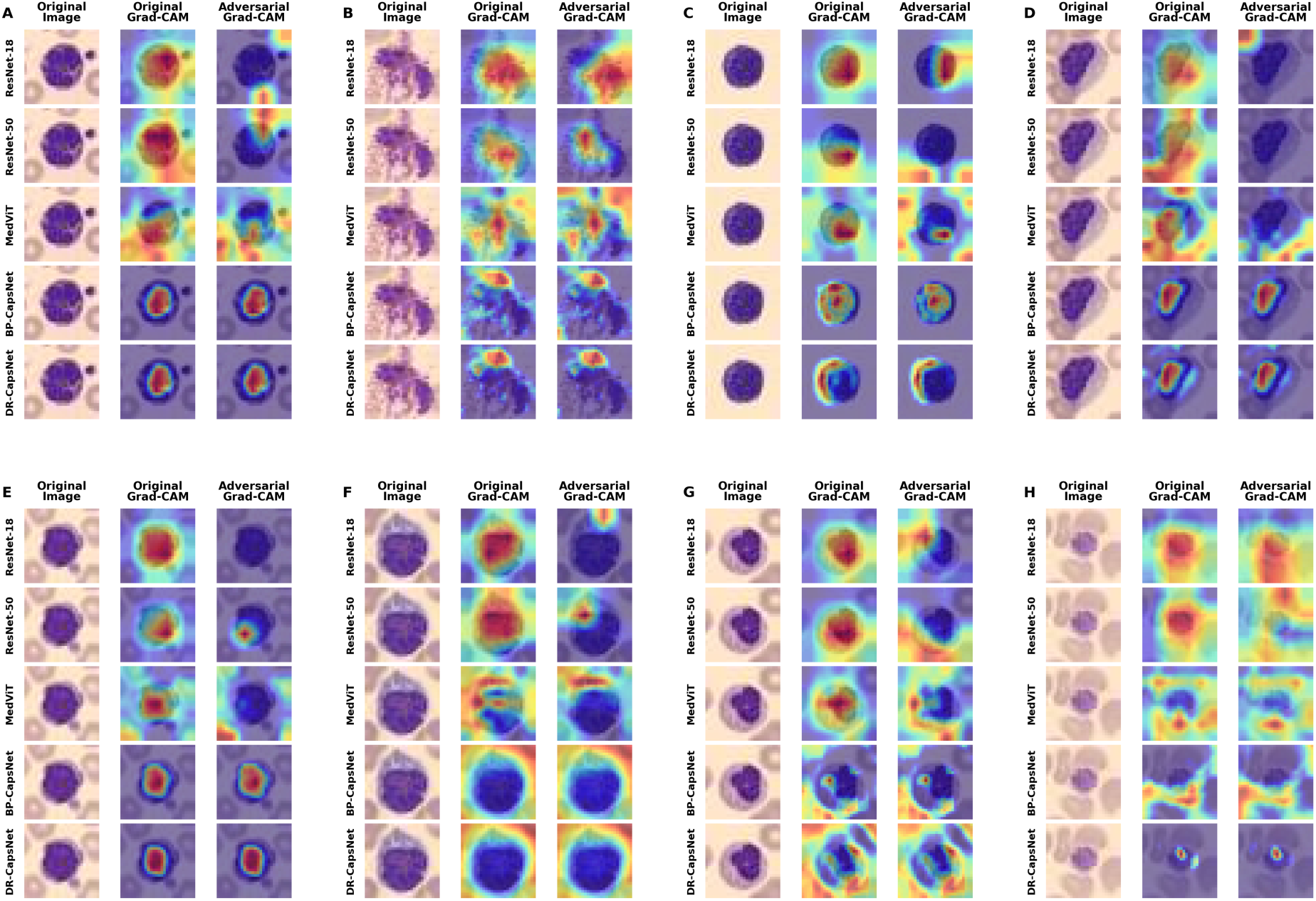
Supplemental Gradient-weighted Class Activation Mapping (Grad-CAM) analysis. Original and adversarial Grad-CAMs with a representative example from each class in the BloodMNIST dataset including a(n) (**A**) basophil, (**B**) eosinophil, (**C**) erythroblast, (**D**) immature granulocyte, (**E**) lymphocyte, (**F**) monocyte (**G**) neutrophil, and (**H**) platelet. CapsNet Grad-CAMs remained consistent after adversarial perturbations, while the Grad-CAMs for other models tended to shift focus.

